# Assessment of the outbreak risk, mapping and infestation behavior of COVID-19: Application of the autoregressive and moving average (ARMA) and polynomial models

**DOI:** 10.1101/2020.04.28.20083998

**Authors:** Hamid Reza Pourghasemi, Soheila Pouyan, Zakariya Farajzadeh, Nitheshnirmal Sadhasivam, Bahram Heidari, Sedigheh Babaei, John P. Tiefenbacher

## Abstract

Infectious disease outbreaks pose a significant threat to human health worldwide. The outbreak of pandemic coronavirus disease 2019 (COVID-2019) has caused a global health emergency. Identification of regions with high risk for COVID-19 outbreak is a major priority of the governmental organizations and epidemiologists worldwide. The aims of the present study were to analyze the risk factors of coronavirus outbreak and identify areas with a high risk of human infection with virus in Fars Province, Iran. A geographic information system (GIS)-based machine learning algorithm (MLA), support vector machine (SVM), was used for the assessment of the outbreak risk of COVID-19 in Fars Province, Iran. The daily observations of infected cases was tested in the third-degree polynomial and the autoregressive and moving average (ARMA) models to examine the patterns of virus infestation in the province and in Iran. The results of disease outbreak in Iran were compared with the data for Iran and the world. Sixteen effective factors including minimum temperature of coldest month (MTCM), maximum temperature of warmest month (MTWM), precipitation in wettest month (PWM), precipitation of driest month (PDM), distance from roads, distance from mosques, distance from hospitals, distance from fuel stations, human footprint, density of cities, distance from bus stations, distance from banks, distance from bakeries, distance from attraction sites, distance from automated teller machines (ATMs), and density of villages – were selected for spatial modelling. The predictive ability of an SVM model was assessed using the receiver operator characteristic – area under the curve (ROC-AUC) validation technique. The validation outcome reveals that SVM achieved an AUC value of 0.786 (March 20), 0.799 (March 29), and 86.6 (April 10) a good prediction of change detection. The growth rate (GR) average for active cases in Fars for a period of 41 days was 1.26, whilst it was 1.13 in country and the world. The results of the third-degree polynomial and ARMA models revealed an increasing trend for GR with an evidence of turning, demonstrating extensive quarantines has been effective. The general trends of virus infestation in Iran and Fars Province were similar, although an explosive growth of the infected cases is expected in the country. The results of this study might assist better programming COVID-19 disease prevention and control and gaining sorts of predictive capability would have wide-ranging benefits.

## Introduction

In December 2019 several pneumonia infected cases were reported in Wuhan, China [1-2]. In January 2020, a novel coronavirus (2019-nCoV) that was later formally named COVID-19 was approved in Wuhan [3]. It was announced that the disease is a severe acute respiratory syndrome coronavirus 2 (SARS-CoV-2). The virus elevated concerns within China as well as the global community as it was believed to be transmitted from human to human [4]. Initially, China witnessed the largest outbreak in Hubei and other nearby provinces. The spread in China was controlled soon thereafter through stringent preventive measures, but other parts of the world (Europe, the Middle East, and the United States) were increasingly affected by the outbreak through transmission by infected travellers from China. A similar outbreak soon followed in other Asian countries [5]. Its global spread to more than 150 countries led to the declaration in mid-March 2020 that COVID-19 was a pandemic [6]. By April 10, 2020, there were nearly 1.70 million cases worldwide with 102684 deaths attributed to COVID-19 [7]. Currently, the United States has the largest number of confirmed cases, while Italy has reported the highest number of casualties [7-8]. Iran with 68,192 recorded cases and 4232 deaths is the most affected country in the Middle East (as of April 10, 2020) and infected cases are expected to surge in the coming days [7, 9]. The outbreak of COVID-19 has disrupted and depressed the world economy, whereas Iran is among the most severely affected by massive economic losses, largely compounded by politically motivated sanctions imposed by other governments [10]. The problem has been exacerbated as no specific medicine is yet realized for COVID-19 disease treatment, though there are a few pre-existing drugs that are being tested, so regions are presently concentrating their efforts on maintaining the infection rate in a level that assists to reduce virus spread [11]. This has led to most states imposing lockdowns, encouraging social distancing, and restricting the sizes of gatherings to limit transmission [12]. There is a pressing necessity for scientific communities to aid governments in their efforts to control and prevent transmission of the virus [13].

During previous virus outbreaks stemming from Zika, influenza, West Nile, Dengue, Chikungunya, Ebola, Marburg, and Nipah, geographic information systems (GISs) have played significant roles in providing significant insight via risk mapping, spatial forecasting, monitoring spatial distributions of supplies, and providing spatial logistics for management [13]. In this current situation, risk mapping is critical and may be used to aid governments’ need for tracking and management of the disease as it spread in places with the highest risk. Sánchez-Vizcaíno et al. [14] used a multi-criteria decision making (MCDM) model to map the risk of Rift Valley fever in Spain. Traditional statistical techniques had been also used to detect the risk of outbreak [14]. Reeves et al. [15] employed an ecological niche modelling (ENM) technique for mapping the transmission risk of MERS-CoV; the Middle Eastern name for the coronavirus known as SARS-CoV-2. Similar techniques have been in the Nyakarahuka et al. [16] study to map Ebola and Marburg viruses risks in Uganda. They assessed the importance of environmental covariates using the maximum entropy model.

More recently, the use of machine learning algorithms (MLAs) for mapping the risk of transmission of viruses has been increasing which is due to the demonstrated superior (and more accurate) predictive abilities of the MLA models over traditional methods [17]. Jiang et al. [18] employed three MLAs – backward propagation neural network (BPNN), gradient boosting machine (GBM), and random forest (RF) – to map the risk of an outbreak of Zika virus. Tien Bui et al. (2019) compared different MLAs – artificial neural network (ANN) and support vector machine (SVM) with ensemble models including adaboost, bagging, and random subspace – for modelling malaria transmission risk. Similarly, GBM, RF, and general additive modelling (GAM) were used by Carvajal et al. [19] to model the patterns of dengue transmission in the Philippines. Mohammadinia et al. [20] employed geographically weighted regression (GWR), generalized linear model (GLM), SVM, and ANN to develop a forecast map of leptospirosis; GWR and SVM produced highly accurate predictions. The literature shows that very few studies have tried to use GIS for analysis of COVID-19 outbreak in human communities. Kamel Boulos and Geraghty [21] described the use of online and mobile GIS for mapping and tracking COVID-19 whilst Zhou et al. [13] revealed the challenges of using GIS for SARS-CoV-2 big data sources. To our knowledge, there has been no study with focus on mapping the outbreak risk of the COVID-19 pandemic. The aims of the present study were to analyze the risk factors of coronavirus outbreak and test the SVM model for mapping areas with a high risk of human infection with virus in Fars Province, Iran. The outcome of the present study lays a foundation for better programming and understanding the factors that accelerate virus spread for use in disease control plans in human communities.

## Materials and methods

### Study area

The study area is in the southern part of Iran with an area of 122608 square kilometres located between 27°2°′ and 31°42′ N and between 50°42 ′ and 55°36′ E. Fars is the fourth largest province in Iran (7.7 % of total area) with a population density of 4851274 (based on in 2016 report). Fars Province is divided into 36 counties, 93 districts, and 112 cities (Fig 1).

**Fig 1.**
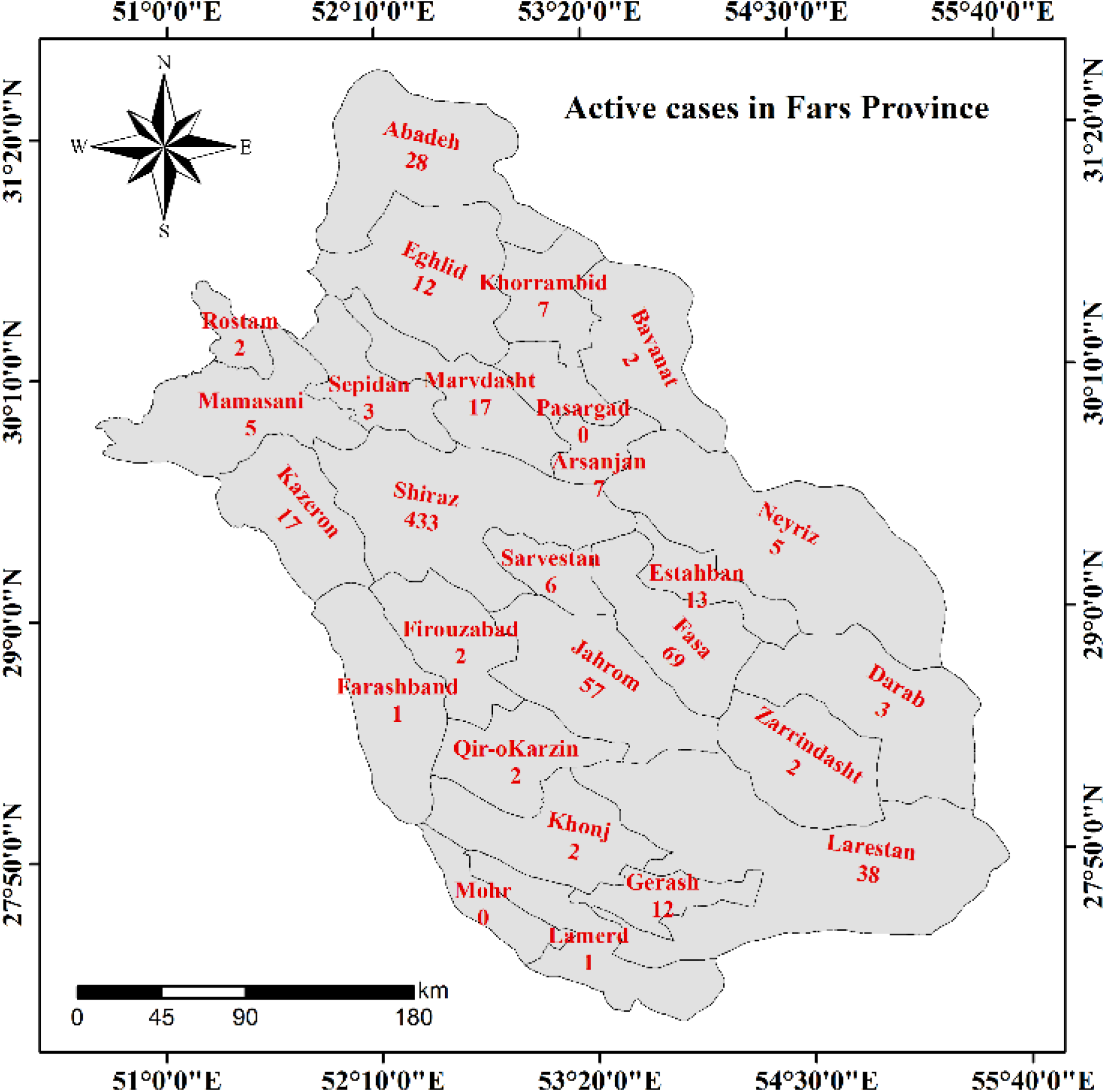
The counties of Fars Province, Iran, and the number of COVID-19 infected case identified from March 29, 2020.

### Methodology

The multi-phased workflow implemented in this investigation (Fig. 2) is described comprehensively below.

**Fig 2.**
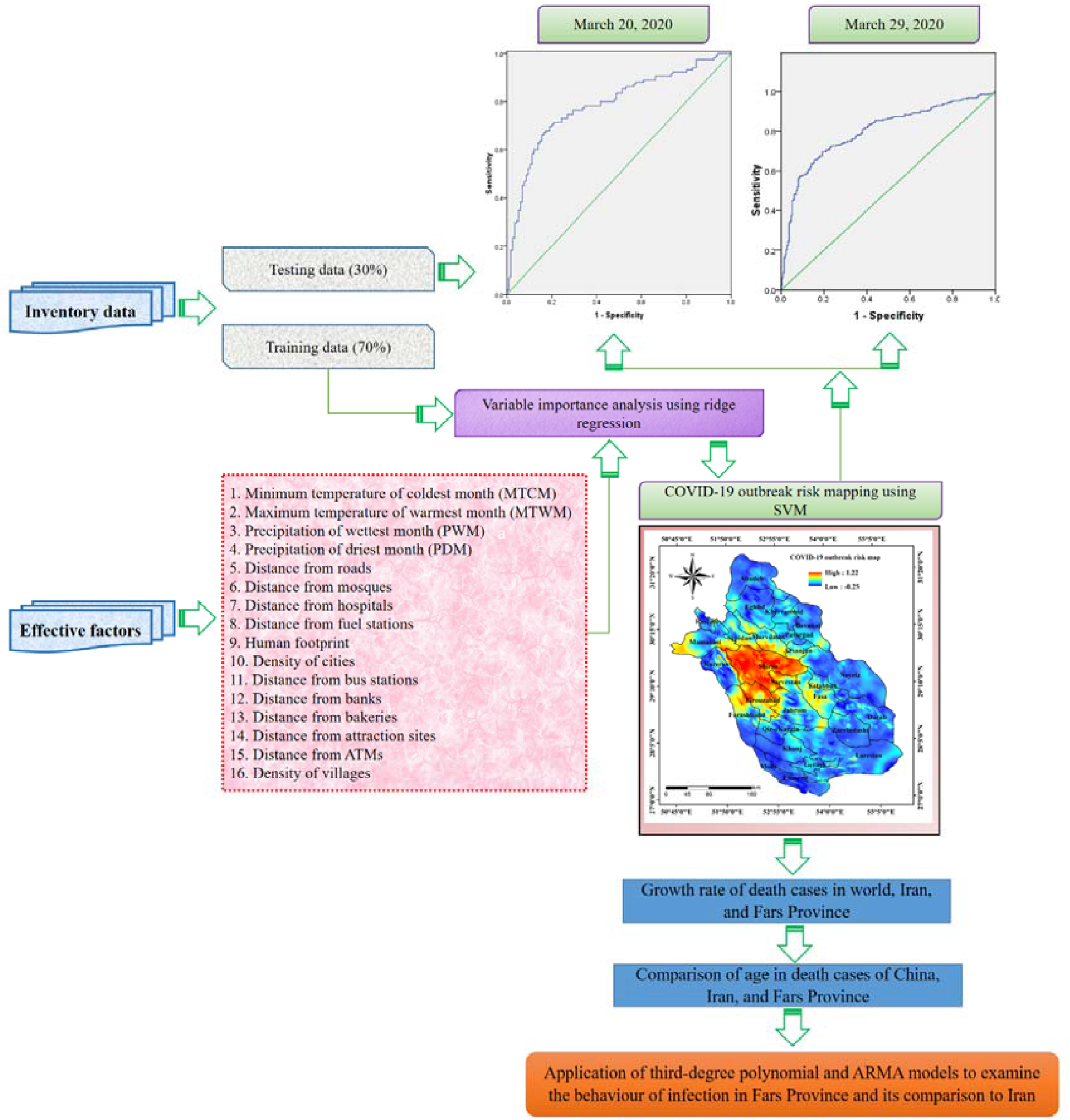
The methodological framework followed in this study.

### Preparation of location of COVID-19 active cases

A dataset of active cases of COVID-19 in Fars was prepared to analyse the relationships between the locations of active cases and the effective factors that may be useful for predicting outbreak risk. The data utilized in this research was collected on April 10, 2020 from Iranian’s Ministry of Health and Medical Education (IMHME).

### Preparation of effective factors

Choosing the appropriate effective factors to predict the risk of pandemic spread is vital as its quality affects the validity of the results [17]. Since, there have been no previous studies of risk for COVID-19 distribution, the selection of effective factors is a quiet challenging task. Ongoing research on the pandemic has revealed that local and community-wide transmission of the virus largely happens in public places where the most people are likely to come into contact with largest number of potential carriers of the infection [22]. Wang et al. [23] indicated that meteorological conditions, such as rapidly warming temperatures in 439 cities around the world resulted in a decline of COVID-19 cases. Accordingly, in this research, we selected sixteen most relevant effective factors for the outbreak risk mapping of COVID-19 in Fars Province of Iran, which includes minimum temperature of coldest month (MTCM), maximum temperature of warmest month (MTWM), precipitation in wettest month (PWM), precipitation of driest month (PDM), distance from roads, distance from mosques, distance from hospitals, distance from fuel stations, human footprint, density of cities, distance from bus stations, distance from banks, distance from bakeries, distance from attraction sites, distance from automated teller machines (ATMs) and density of villages. All the effective factors employed in this research are generated using the ArcGIS 10.7.

A few studies have established that variation in temperature would impact the transmission of COVID-19 [23]. It has been also reported that alteration in temperature would have impacted the SARS outbreak, which was caused by the identical type of coronavirus as SARS-CoV-2 [24]. Recently, Ma et al. [2] disclosed that surge in temperature and humidity conditions have resulted in the decline of death caused by SARS-CoV-2. Thus, climatic factors such as temperature and precipitation can have an impact in the outbreak of SARS-CoV-2. The temperature and precipitation data namely MTWM, MTCM, PDM and PCM of Fars Province is acquired from world climatic data (https://www.worldclim.org/). In this study, the MTWM of the Fars Province ranges from 27.7°C to 41.8°C (Fig 3a) whereas MTCM ranges between −15.3°C and 10.4°C (Fig 3b). The PWM of the study area varies between 28 mm and 86 mm (Fig 3c) and also the PDM is presented in Fig 3d.

**Fig 3.**
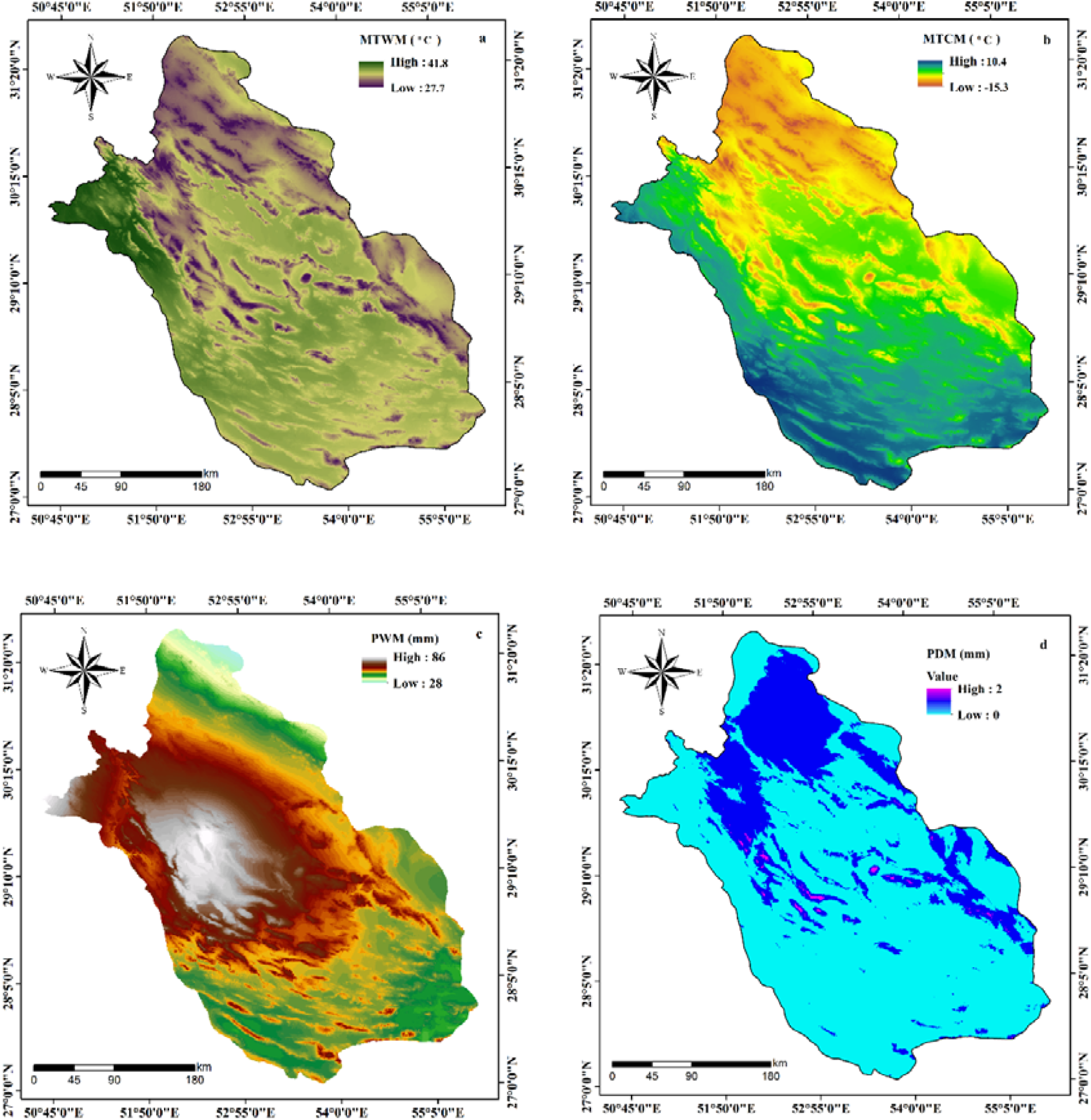
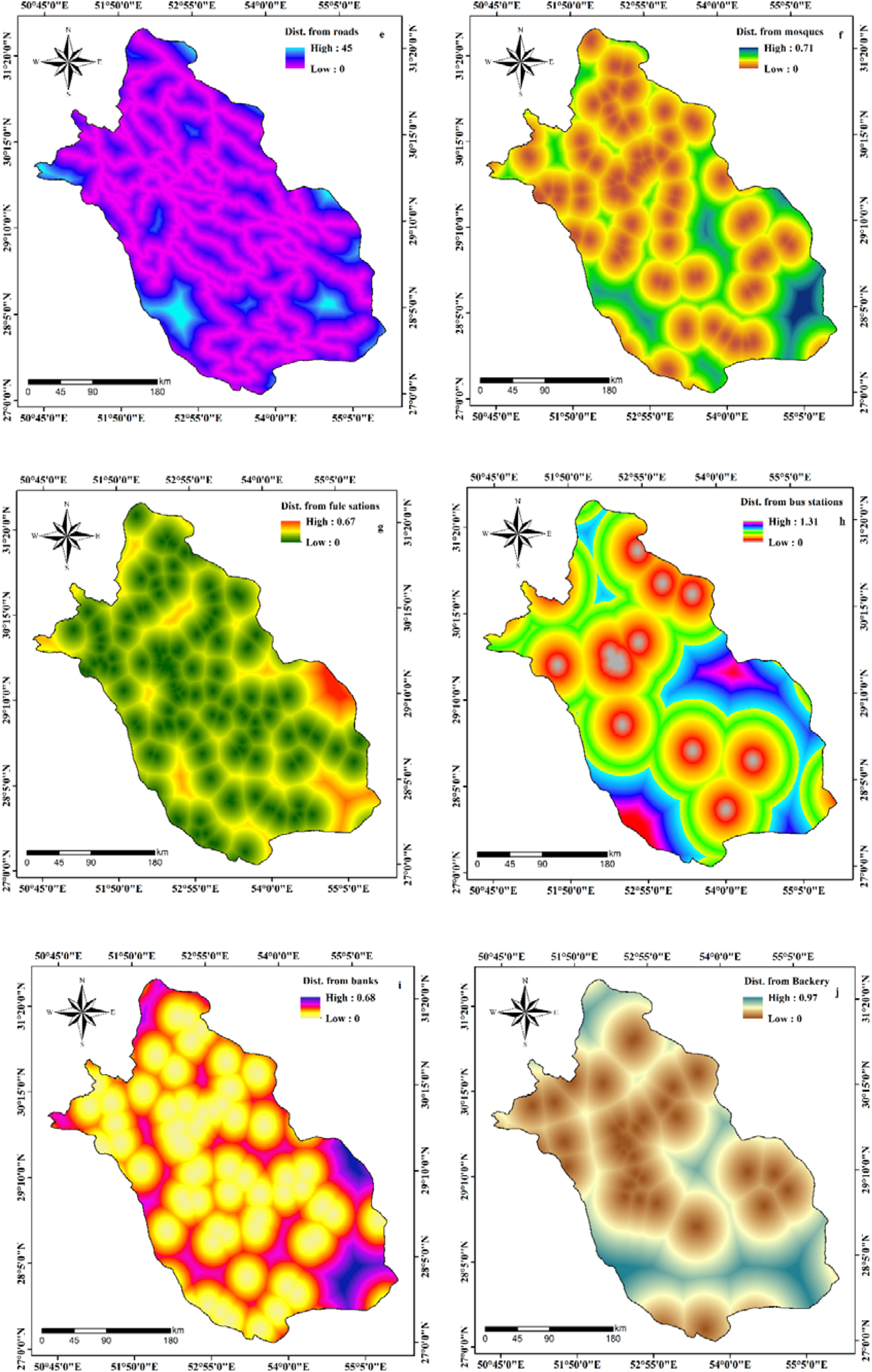
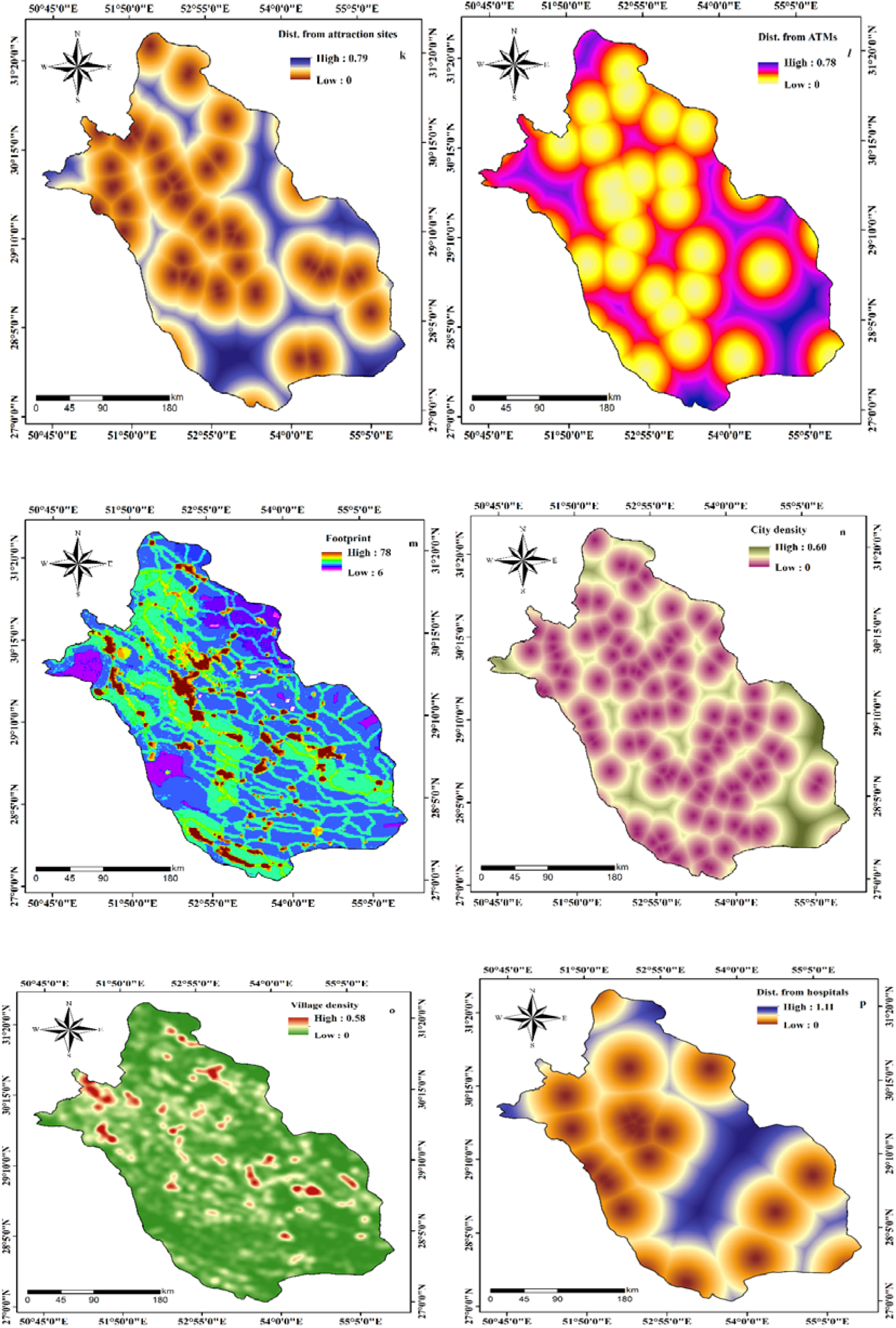
Preparation of effective factors of COVID-19 outbreak

The proximity to various public places including roads, mosques, hospitals, fuel stations, bus stations, banks, bakeries, attraction sites, and ATMs where people come in close contact to each other can also be considered as significant factors that influence the distribution of COVID-19. The distance from roads ranges from 0 to 45 in the study area (Fig 3e) whereas the distance from mosques varies between 0 and 0.71 (Fig 3f) and the distance from fuel stations spans 0 to 0.67 (Fig 3g). The distance from bus stations, banks, bakeries, attraction sites, and ATMs of Fars Province have the minimum value of 0 and maximum value of 1.31, 0.68, 0.97, 0.79, and 0.78 respectively (Fig 3h – 3l). Since, humans are the potential carriers of the COVID-19, the use of human footprint (HFP) can aid in understanding the terrestrial biomes on which humans have more influence and access [25]. In this study, HFP of the study area is acquired from the Global Human Footprint Dataset. The HFP of Fars Province ranges from 6 to 78 (Fig 3m) where the minimum value represents the places having least access by humans and the maximum value refers to those regions having highest human influence and access. The density of population is also considered to be an important factor for the spread of the disease [26-27]. Gilbert et al., [28] revealed that the number of COVID-19 cases were proportional to the population density in Africa. Accordingly, in this research, density of cities and villages were assessed and the outcome displays that density of cities in Fars Province ranges between 0 and 0.60 (Fig 3n) while the density of villages varies from 0 to 0.58 (Fig 3o). The distance from hospitals ranged from 0 to 1.11 (Fig 3p).

### Evaluation of variable importance using ridge regression

The association among the location of COVID-19 active cases and effective factors were evaluated using ridge regression in order to assess the significance of individual effective factor in predicting the outbreak risk [17]. To our knowledge, no previous study in epidemic outbreak risk mapping have utilized ridge regression in determining the significance of effective factors. However, the ridge regression algorithm has been utilized for modelling purposes in various fields [29]. It was first given by Hoerl and Kennard [30] which exploits L_2_ norm of regularization for lessening the model complication and controlling overfitting. Ridge regression was also developed to avoid the excessive instability and collinearity problem caused by least square estimator [31]. The ‘caret’ package (https://cran.r-project.org/web/packages/caret/caret.pdf) of R 3.5.3 was utilized for assessing the variable importance using ridge regression.

### Machine learning algorithm (MLA)

#### Support vector machine

SVM is an extensively exercised MLA in diverse fields of research that functions on the principle of statistical learning concept and structural risk minimization given by Vapnik [32], which is utilized for classification as well as regression intricacies [33-34]. SVM has a high efficacy in classifying both linearly separable and inseparable data classes [35]. It utilizes an optimal hyperplane to distinguish linearly divisible data whereas kernel functions are employed for transforming inseparable data into a higher dimensional space so that it can be easy categorized [36]. Assume a calibration dataset to be (s_m_, t_m_), where m is 1, 2, 3…, x; s_m_ refers to the sixteen independent factors; t_m_ denotes 0 and 1 that resembles risk and non-risk classes and × represents the total amount of calibration data. This algorithm tries to obtain an optimal hyperplane for classifying the aforementioned classes by utilizing the distance between them, which can be formulated as follows [37]:

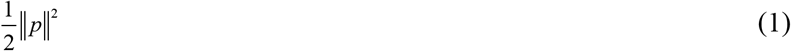

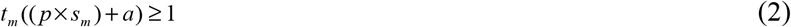

where, ‖*p* ‖ denotes the rule of normal hyperplane; a refers to a constant. When Lagrangian multiplier (*λ*_*m*_) and cost function is introduced, the expression can be given as follows [38]:

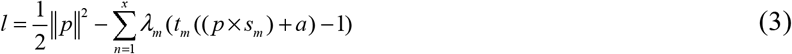

In case of inseparable dataset, a slack covariate *δ*_*m*_ is added into the previous Eq. (2) that is provided as follows [32]:

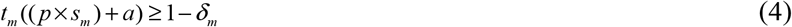

Accordingly, the Eq. (3) can be described as follows [32]:

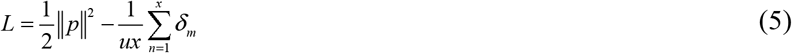

Moreover, SVM contains four kernel functions (linear, polynomial, radial basis function: RBF and sigmoid) for making an optimal margin in case of inseparable dataset [32]. Mohammadinia et al. [20] revealed that RBF kernel type produces high prediction accuracy than other kernel types for epidemic outbreak risk mapping. Thus, in this study, RBF is used for creating decision boundaries and the kernel function is expressed as follows [32]:

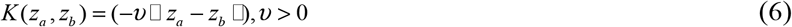

where, K(z_a_, z_b_) refers to kernel function and *v* represents its parameter.

### Analysis of growth rate for active and death cases of COVID-19

In this study, the growth rate (GR) of active and death cases around the world, Iran, and Fars Province were evaluated using the data acquired from WHO and IMHME between February 26, 2020 and April 10, 2020 for active cases and from March 3, 2020 to April 10, 2020 for death cases.

### Validation of outbreak risk map

The cross-checking of calibrated model using untouched testing data is vital for determining the scientific robustness of the prediction [33]. In this research, we utilized ROC-AUC curve values for the validation of COVID-19 outbreak risk map generated using SVM model. It is a widely utilized validation technique for analysing the predictive ability of a model [35]. A model is determined to be perfect, very good, good, moderate and poor if the AUC values were 1.0-0.9, 0.9-0.8, 0.8-0.7, 0.7-0.6 and 0.6-0.5 respectively [39].

### Models for infection cases trend

The behavior of the variable infection cases was captured by a third-degree polynomial or cubic specification as follows:

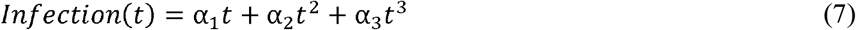

Where *Infection*(*t*) represents the total infected cases in day t and t denotes the days starting from 19th of February for Iran and one week later for Fars province. Also, other specifications including quadratic as well as fourth-degree polynomial specifications were examined and based on the predictions, the cubic form was selected against other specifications. In the literature, this form of the specification has been applied by Aik et al. [40] to examine the Salmonellosis incidence in Singapore. We also used an ARMA model to compare the process generating the variable for Iran and Fars province. This model includes two processes: Autoregressive (AR) and Moving Average (MA) process. An ARMA model of order (p,q) can be written as [41]:

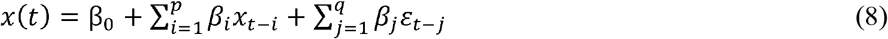

Where × is the dependent variable and *ε* is the white noise stochastic error term. In the applied model, × shows the total infected cases and t is the days starting from the first day of happening infection cases. Benvenuto et al. [42] also applied an ARIMA model to predict the epidemiological trend of COVID-2019.

## Results

### Outcome of the variable importance analysis

The analysis of variable importance using ridge regression revealed that distance from bus stations, distance from hospitals, and distance from bakeries have the highest significance whereas distance from ATMs, distance from attraction sites, distance from fuel stations, distance from mosques, distance from road, MTCM, density of cities and density of villages exhibit moderate importance. The effective factors such as distance from banks, MTWM, HFP, PWM and PDM were the least influential factors (Fig 4).

**Fig 4.**
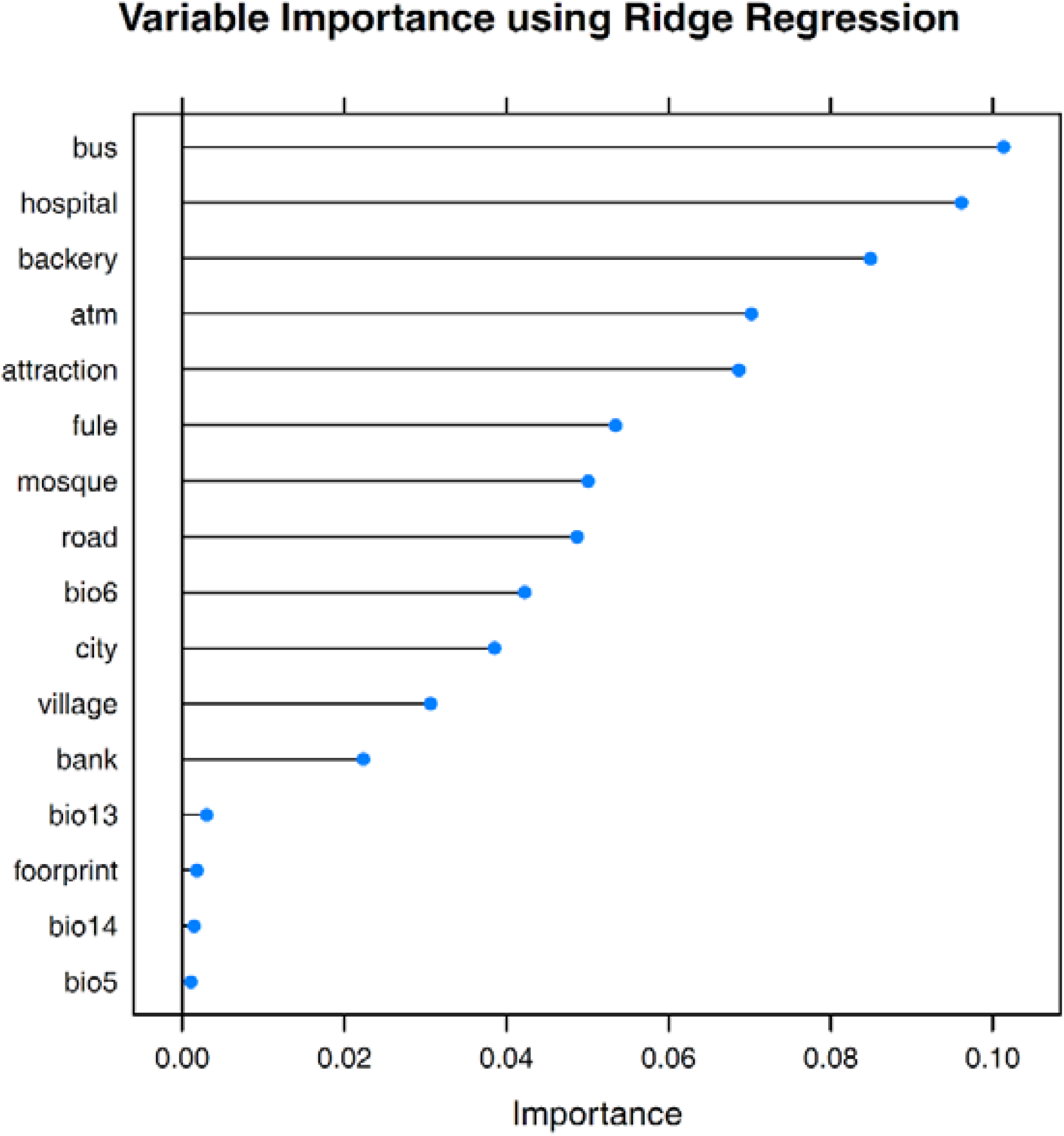
Variable importance of each effective factors (bus: distance from bus stations; hospital: distance from hospitals; bakery: distance from bakeries; atm: distance from ATMs; attraction: distance from attraction sites; fuel: distance from fuel stations; mosque: distance from mosques; road: distance from road; bio6: MTCM; city: density of cities; village: density of villages; bank: distance from banks; bio13: MTWM; footprint: HFP, bio14: PWM; bio5: PDM.

### COVID-19 outbreak risk map using SVM

The COVID-19 outbreak risk map generated using SVM displays that risk of SARS-CoV-2 ranges from −0.25 to 1.22 (March 29) and −0.35 to 1.21 (April 10) where −0.25 and −0.35 represents the lower risk of SARS-CoV-2 outbreak and 1.22 and 1.21 indicates the regions of Fars Province which is likely to experience a higher risk of COVID-19 outbreak (Fig 5, a-b). It can be observed from Fig 5b (April 10) that Shiraz County and its surrounding counties including Firouzabad, Jahrom, Sarvestan, Arsanjan, Marvdasht, Sepidan, Abadeh, Khorrambid, Rostam, Larestan and Kazeron of Fars Province has the highest risk of being the epicentre of SARS-CoV-2 outbreak. Apart from which counties like Eghlid, and Fasa also lie in the high risk zone.

**Fig 5.**
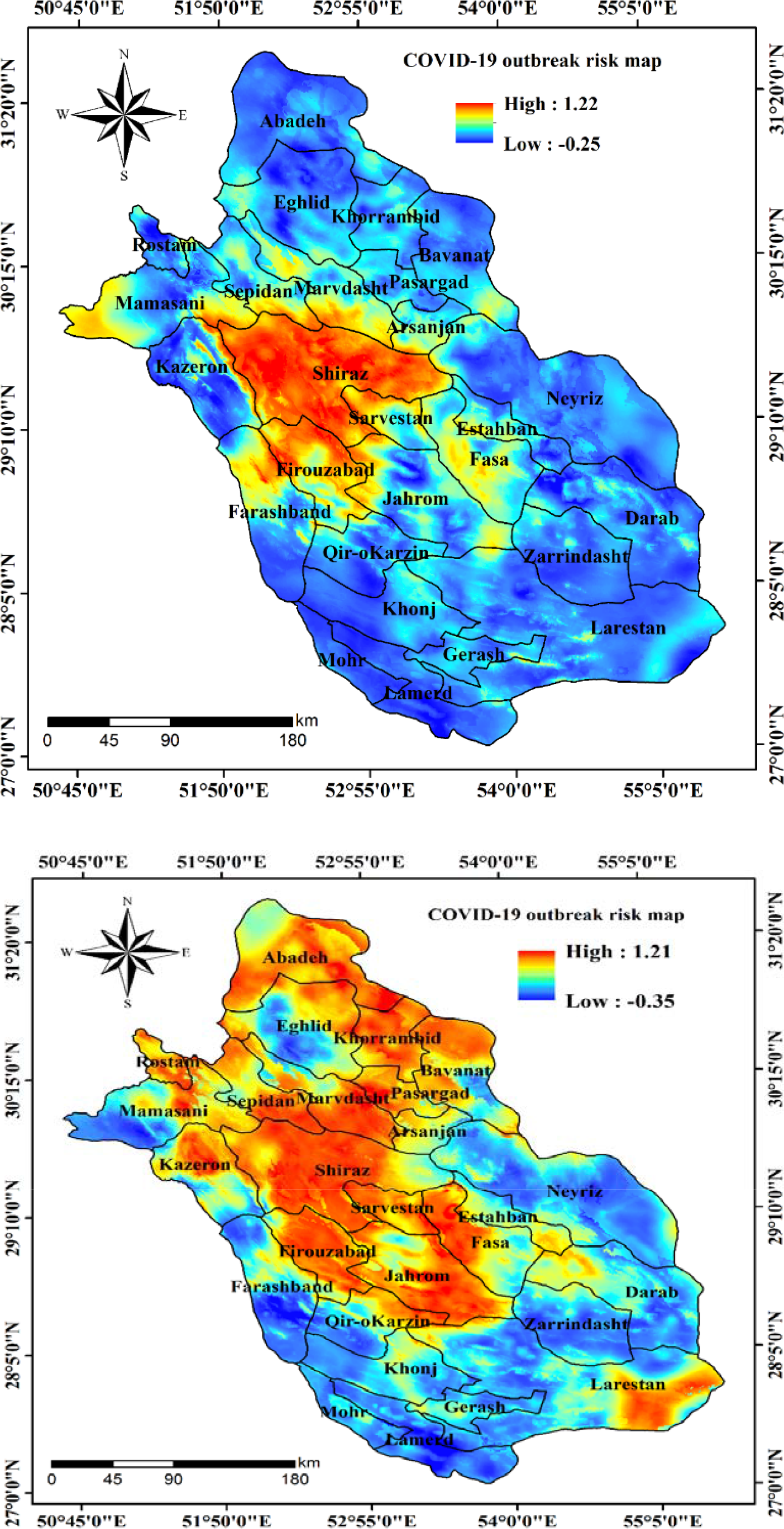
The COVID-19 outbreak risk map a) on March 29, 2020 and b) on April 10, 2020

### Outcome of growth rate analysis

The results of GR of active cases in world, Iran, and Fars Province are presented in Fig 6. Our results displayed that the highest active cases in world, Iran, and Fars Province was related to March 11 (GR=1.95), Feb 26 (GR=2.41), and March 15 (GR=4.8), respectively. Also, the outcome stated that GR average of active cases in world, Iran, and Fars Province reported since March 1 to April 10 was 1.13, 1.13, and 1.25, respectively. Our observations demonstrated that the highest GR of active cases in Fars Province was on March 16 (GR=4.80), March 09 (GR=3.20), March 20 (GR=2.40), March 22 (GR=2.10), April 1^st^ (GR=2.10), and March 26 (GR=1.90). On the other hand, the analyses indicated that between February 27 and February 29, the GR of active cases was zero in Fars Province, followed by a GR value of 0.3 in March 14, March 19, and March 21, whereas the lowest GR of active cases in world and Iran observed on March 4 (GR=0.89) and March 3 (GR=0.67) respectively.

**Fig 6.**
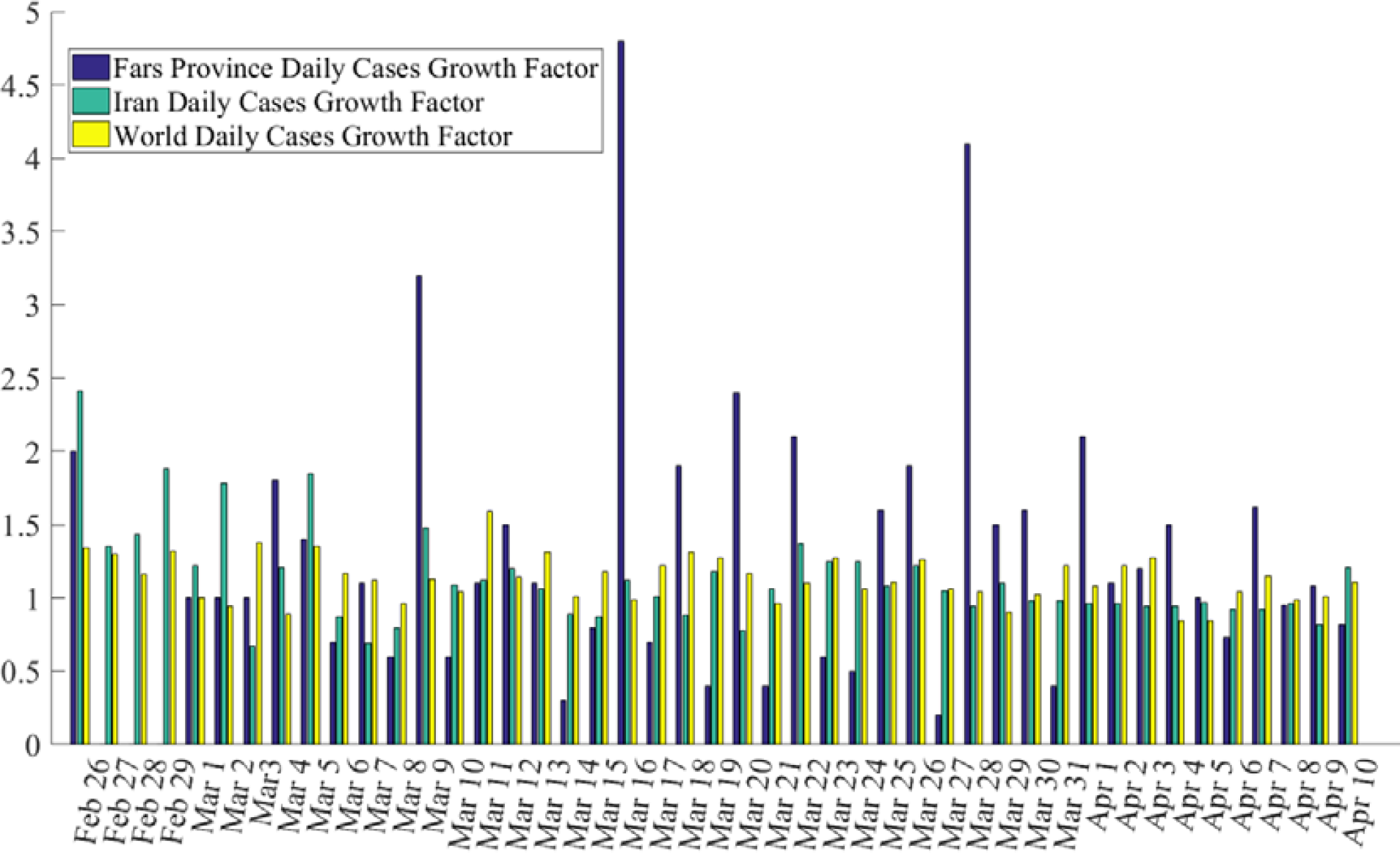
Growth rate of active cases in world, Iran, and Fars Province

Results of death cases in world, Iran, and Fars Province are given in Fig 7.

**Fig 7.**
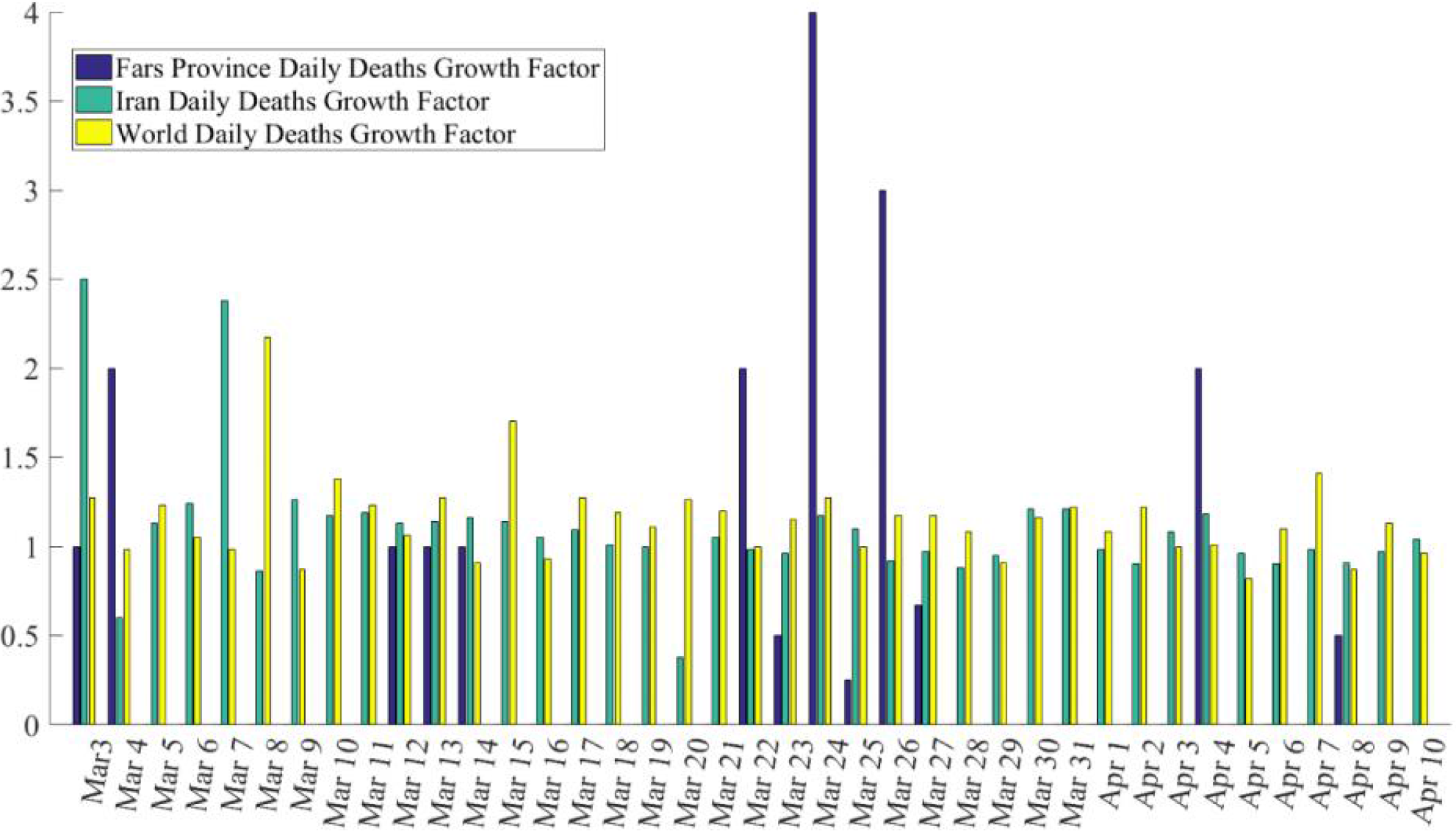
Growth rate of death cases in world, Iran, and Fars Province

In total of 1762 active cases of COVID-19 in Fars Province, 42 died between February 24 and April 10. The highest GR of death cases in Fars Province was reported on March 24 (GR=4.00), March 26 (GR=3.00), March 22 (GR=2.00), March 4 (GR=2.00), and April 5 (GR= 2.00). Our analyses showed that since March 5 to March 11, March 15 to March 21, March 28 to April 4, and April 5 to April 8, the GR of death cases was equal to zero. Although the deaths on March 31, April 3, April 7, and April 10 were 3, 2, 4, and 1, respectively, the daily growth rate is zero. Also, average of the GR in Fars Province during 41 days was 0.49, whereas this rate in world and Iran was observed as 1.15 and 1.10, respectively. Fig 7 shows that the highest GR of death cases in world and Iran was nearly equal during March 08 (GR=2.17) and March 03 (GR=2.50). In contrast, the lowest rate of death case was observed on March 09 (GR=0.87), April 08 (GR=0.87), and March 04 (GR=0.60).

Results of active cases in 31 provinces of Iran country by March 25 is presented in Fig 8. Observations indicate that the number of active cases in the 100,000 population vary from 0.4 to 13.1. This figure also shows that provinces of Bushehr and Fars have the lowest cumulative rate of active cases, whereas the highest rate was observed in Qom, Semnan, Mazandaran, Gilan, and Golestan. The Qom Province was the first place in Iran where the outbreak of COVID-19 was recorded.

**Fig 8.**
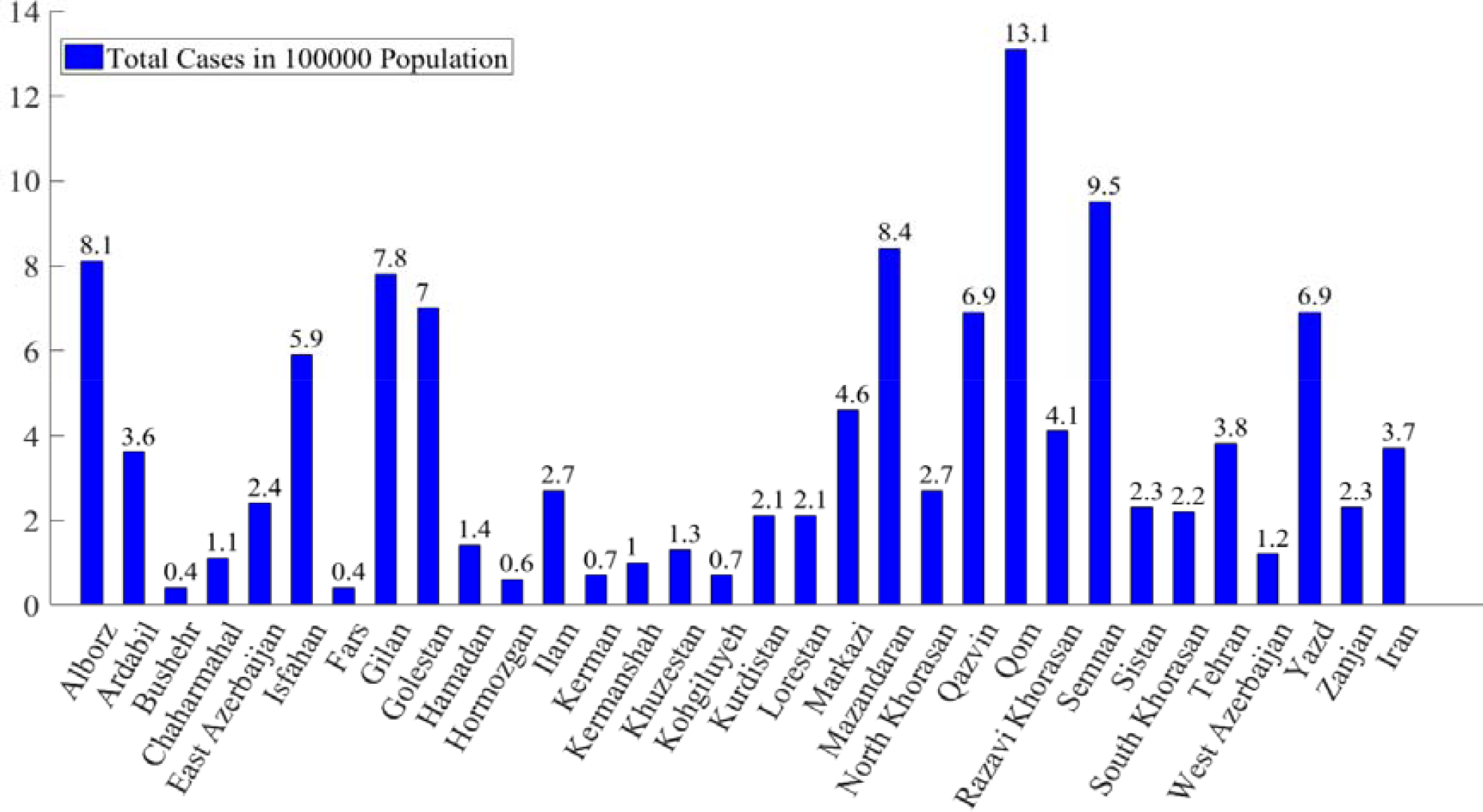
Results of active cases in 31 provinces of Iran country by March 25, 2020

A comparison among age class of death cases in China, Iran, and Fars Province is presented in Table 1. Percentage of death cases in China was related to February 29, whereas for Iran and Fars Province it is related to March 14 and March 31, respectively. Following Table 1 show that age class > 50 years old lie in the highest class of death rate. So, this age class of above 50 years is highly sensitive to COVID-19.

**Table 1.**
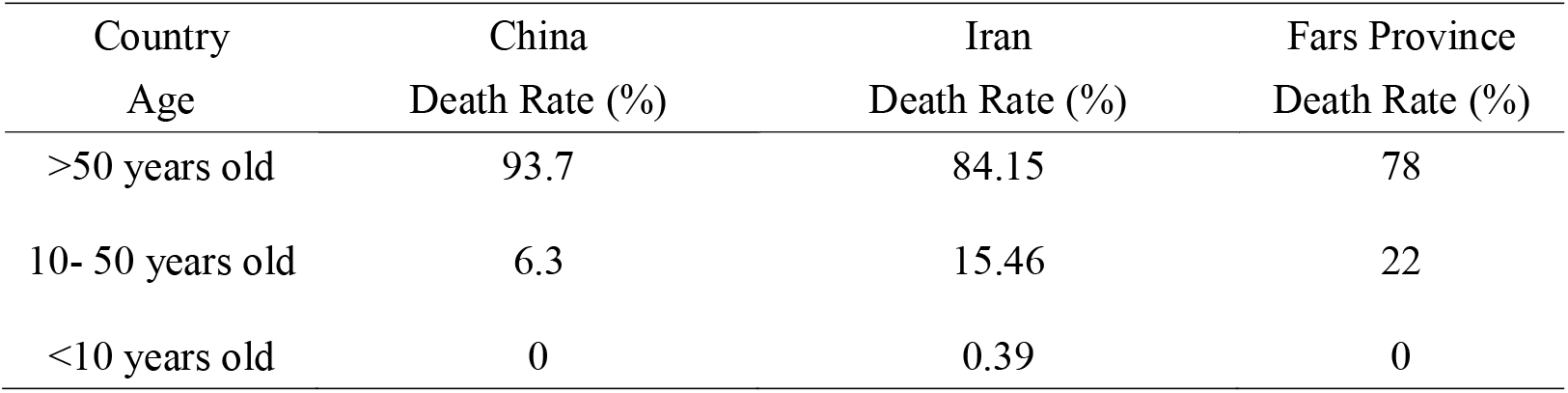
Comparison of age in death cases of China, Iran, and Fars Province

| Country<br>Age | China<br>Death Rate (%) | Iran<br>Death Rate (%) | Fars Province<br>Death Rate (%) |
| --- | --- | --- | --- |
| >50 years old | 93.7 | 84.15 | 78 |
| 10- 50 years old | 6.3 | 15.46 | 22 |
| <10 years old | 0 | 0.39 | 0 |

### Validation outcome of outbreak risk map

The ROC-AUC curve cross-validation technique is utilized in this research for validating the COVID-19 outbreak risk map generated by SVM. The model achieved an AUC value of 0.786 and a standard error of 0.031 indicating a good predictive accuracy when cross-verified using the remaining 30% testing dataset collected on March 20, 2020 (Fig 9 and Table 2).

**Table 2.**
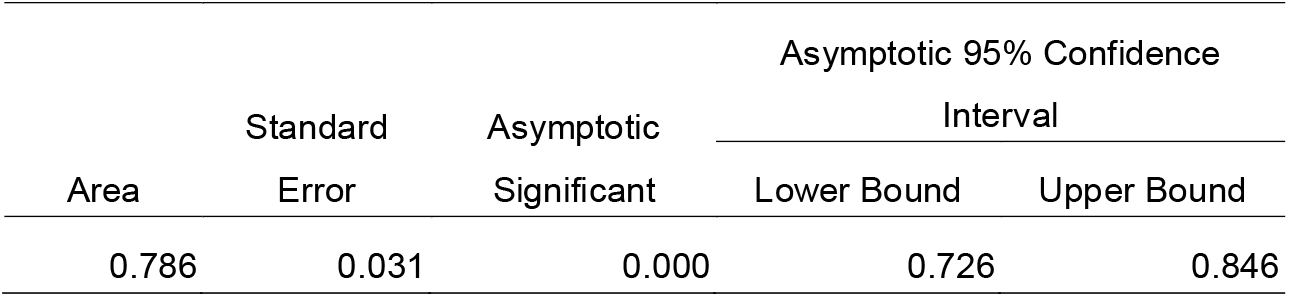
Area under the curve based on data from March 20, 2020

| Area | Standard<br>Error | Asymptotic<br>Significant | Asymptotic 95% Confidence<br>Interval |  |
| --- | --- | --- | --- | --- |
|  |  |  | Lower Bound | Upper Bound |
| 0.786 | 0.031 | 0.000 | 0.726 | 0.846 |

**Fig 9.**
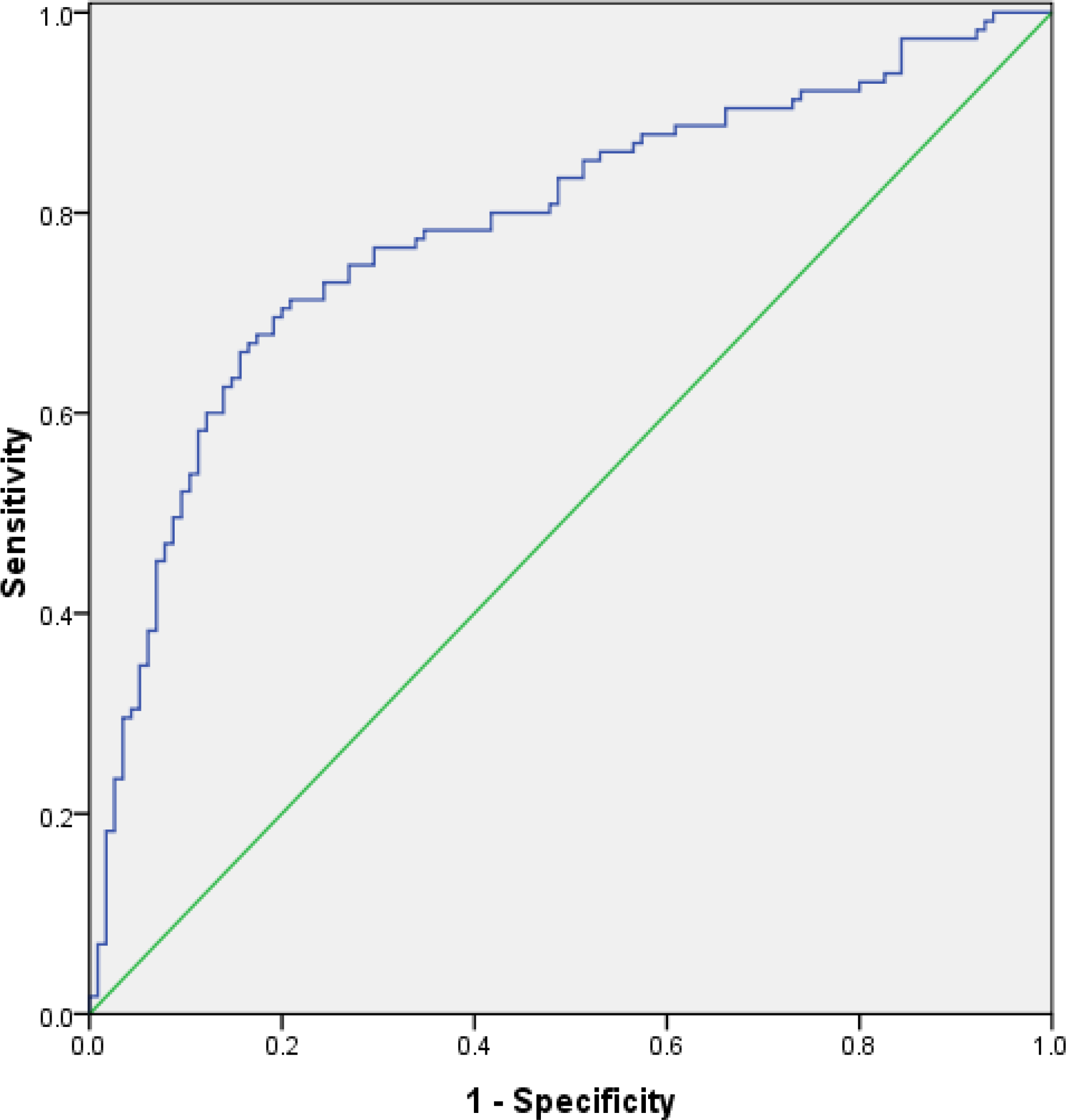
Receiver operator characteristic (ROC) curve based on testing data from March 20, 2020

When tested with active case locations on March 29, 2020, the model achieved an increased AUC value of 0.799 which proves the stable and good forecast precision of the outbreak risk map (Fig 10 and Table 3). Also, change detection on April 10, 2020 show that accuracy of the built models is increased to 86.6% (AUC=0.868) (Fig 11 and Table 4).

**Table 3.** Area under the curve based on data from March 29, 2020

| Area | Standard Error | Asymptotic Significant | Asymptotic 95% Confidence Interval |  |
| --- | --- | --- | --- | --- |
|  |  |  | Lower Bound | Upper Bound |
| 0.799 | 0.022 | 0.000 | 0.756 | 0.841 |

**Table 4.** Area under the curve based on data from April 10, 2020

| Area | Standard Error | Asymptotic Significant | Asymptotic 95% Confidence Interval |  |
| --- | --- | --- | --- | --- |
|  |  |  | Lower Bound | Upper Bound |
| .868 | .015 | .000 | .838 | .898 |

**Fig 10.**
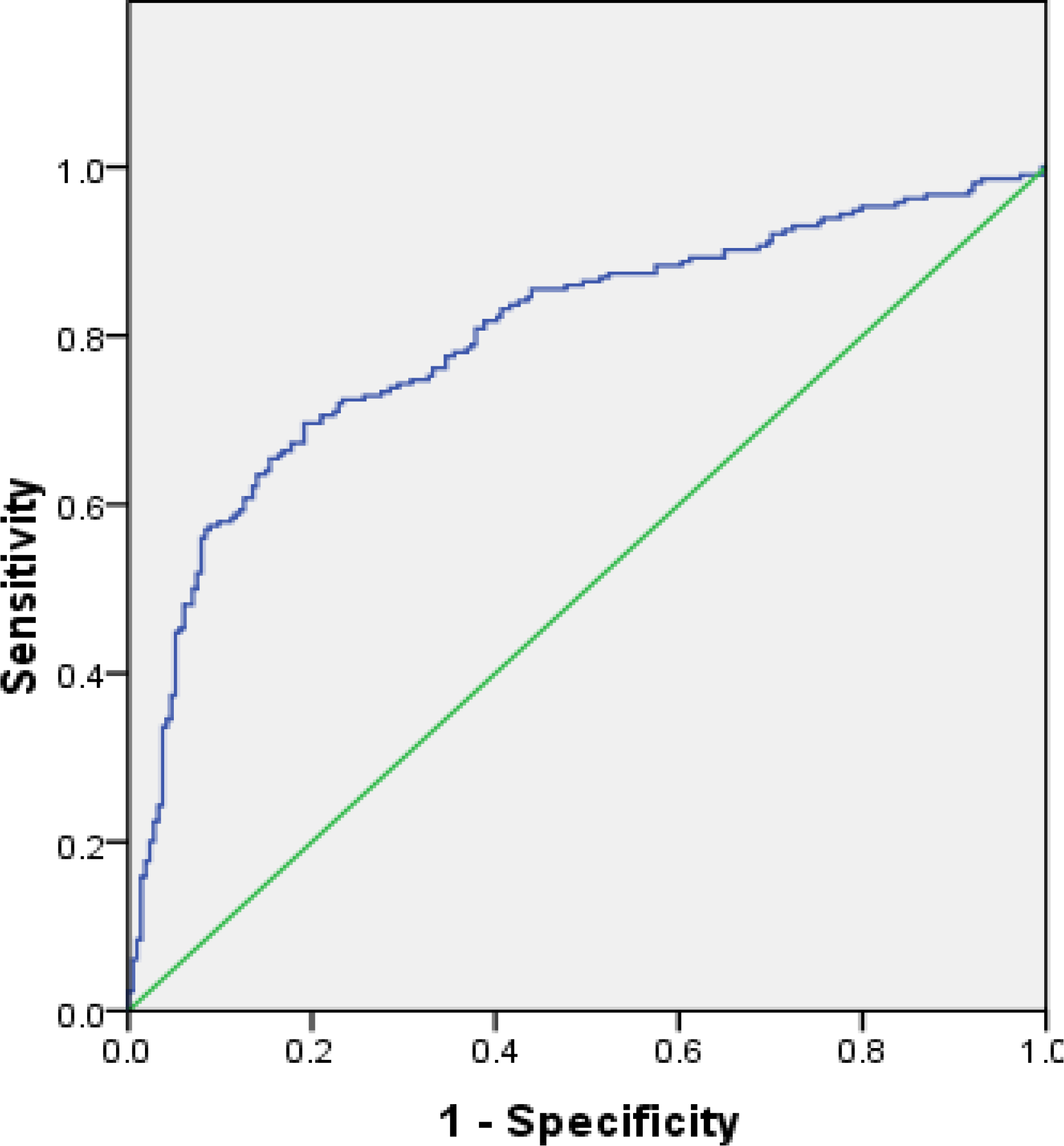
Receiver operator characteristic (ROC) curve based on data from March 29, 2020

**Fig 11.**
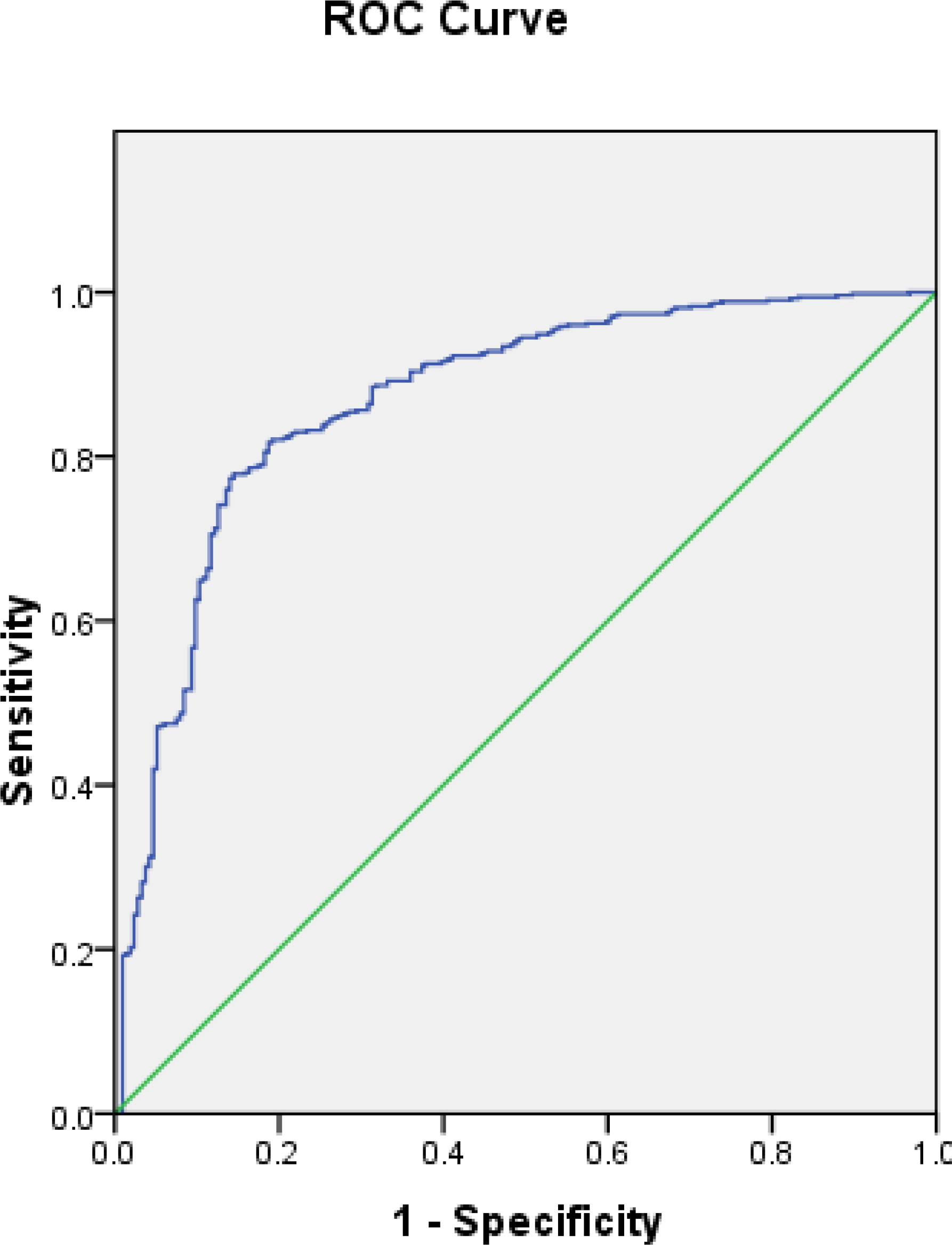
Receiver operator characteristic (ROC) curve based on data from April 10, 2020

### Comparison of Fars province and Iran infection cases

Two tools have been applied to compare the general trend of infection in Fars province and Iran. The first one is a third-degree polynomial model that is presented in Fig 12. Another quantitative model is an ARMA presented in Table 5. Fig 12 shows the trend of infection cases in Iran and Fars province, where predicted values extraordinarily keep pace with the actual values. R ^2^ values also indicate that estimated models have significant predictive power. The infection cases are increasing over the selected horizon.

**Table 5.** The results of autoregressive and moving average (ARMA) model for COVID-19 infection cases of Fars province and Iran

|  | Regressor | Coefficient | Standard error | t-statistics | probability |
| --- | --- | --- | --- | --- | --- |
| Fars province | Constant | 596.015 | 478.41 | 1.24 | 0.221 |
|  | AR(1) | 1.432 | 0.056 | 25.33 | 0.000 |
|  | AR(3) | -0.438 | 0.056 | -7.79 | 0.000 |
|  | Ma(4) | 0.474 | 0.122 | 3.89 | 0.000 |
|  | Adjusted R<sup>2</sup> | 0.997 |  |  |  |
|  | Q(1)a | 1.653 |  |  | 0.199 |
|  | Q(2)a | 1.875 |  |  | 0.392 |
|  | Heteroskedasticity (ARCH) | 0.832 |  |  | 0.367 |
|  | Jarque Berra | 1.083 |  |  | 0.581 |
|  | Inverted AR roots | 0.99 |  |  |  |
|  | Inverted Ma roots | -0.85 |  |  |  |
| Iran | Constant | 43360.05 | 32082.85 | 1.35 | 0.184 |
|  | AR(1) | 1.489 | 0.015 | 99.48 | 0.000 |
|  | AR(3) | -0.492 | 0.014 | -35.01 | 0.000 |
|  | MA(1) | 0.848 | 0.075 | 4.16 | 0.000 |
|  | Adjusted R<sup>2</sup> | 0.999 |  |  |  |
|  | Q(1)a | 1.169 |  |  | 0.194 |
|  | Q(2)a | 1.785 |  |  | 0.410 |
|  | Heteroskedasticity (ARCH) | 1.149 |  |  | 0.289 |
|  | Jarque Berra | 0.088 |  |  | 0.956 |
|  | Inverted AR roots | 0.96 |  |  |  |
|  | Inverted Ma roots | 0.59 |  |  |  |

a<sub>Q(p)</sub> is the significance level of the Ljung–Box statistics in which the first p of the residual autocorrelations are jointly equal to zero.

**Fig 12.**
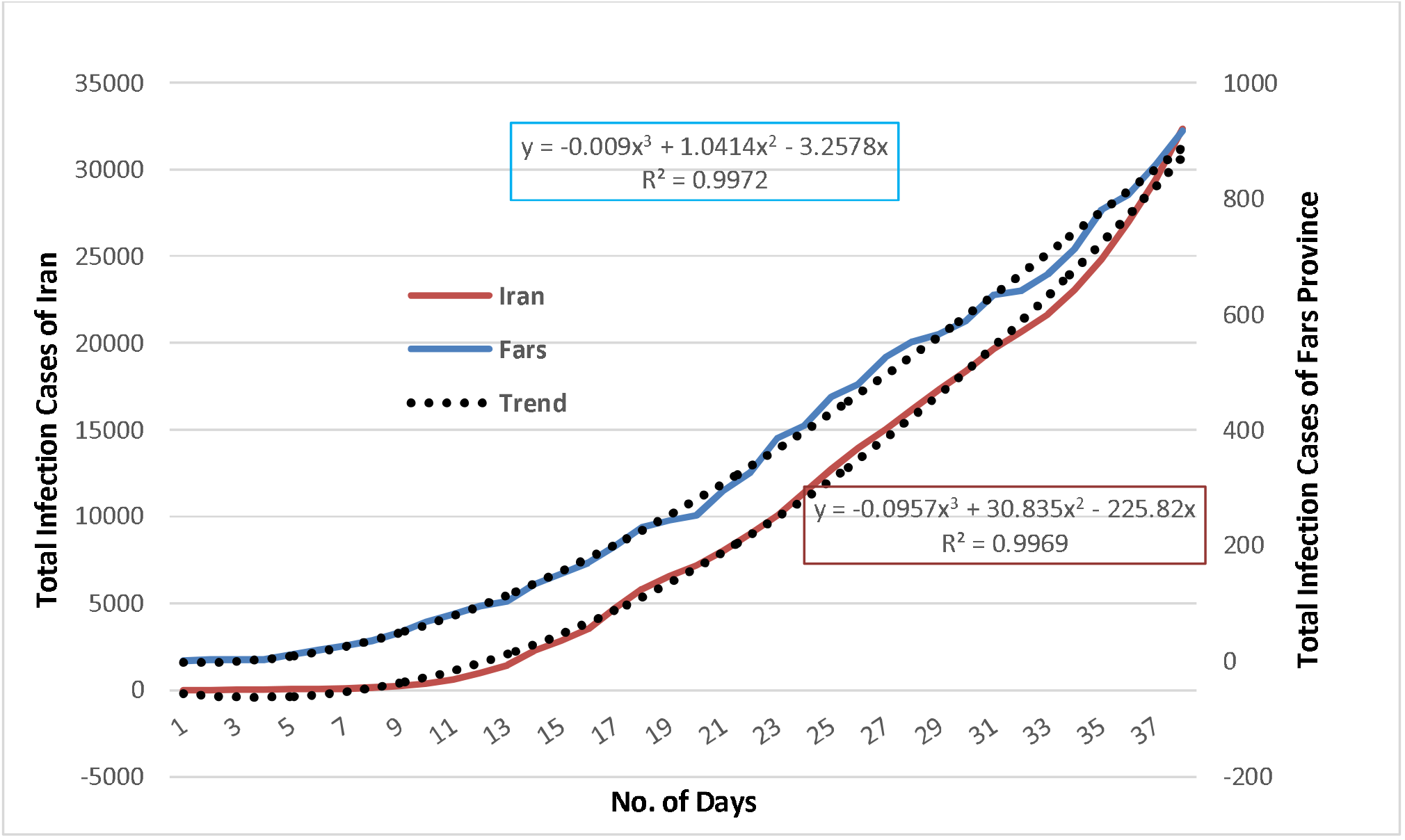
Actual cases versus estimated cases in Fars province and Iran

The first derivative of the estimated model which turns it to a second-degree polynomial equation, represents the daily infection cases. Based on the daily infection model, there is a turning point for both Iran and provincial cases. It was found that the turning point for provincial daily infection is 75. In other words, after 75 days the decreasing trend in the daily infection is expected.

The corresponding value for Iran is 211 that is much higher than the provincial one. There are some evidences showing that a turning point in infection is expected. For instance, it has been reported for SARS incidence [43], HAV [44], ARI [45], and for A (H1N1)v [46]. It is worth noting that a turning point means that after passing the peak it is expected to show a deceasing trend. In the 38^th^ day of infection, Fars province accounts for around 2.84% of the total Iranian cases while its population share is more than 6% (Statistical Center of Iran, 2016). Regarding the values obtained for turning points and the infection share, the measures taken by the provincial government may be considered more effective than those taken in other provinces as a whole. However, it should be taken into consideration that Fars province experienced its first infection cases 7 days after Qom and Tehran, provinces that are considered as starting point for virus outbreak in Iran. This might have given the provincial governmental body and the households to take measures to cope with the widespread outbreak. It is worth noting that the comparison of the specified models is more appropriate to investigate the effectiveness of the measures taken by the corresponding health body rather than using it to predict the future values.

The ARMA time series models for infection variables of the Fars province and Iran are presented in Table 5. These models may show the generating process of the variables in time horizon. It is worth noting that in order to have more comparable models, a 38-day time horizon is selected. This is the period of time that data are available, staring on 19th of February for Iran and one week later for Fars province. As shown in Table 4, the both series are generated by an ARMA (2, 1) process. However, the absolute values of the AR terms for Fars province are lower than those of Iran, indicating a slower process of increasing trend for Fars province compared to those of Iran. However, regarding the values for AR roots, the autoregressive (AR) process for both models isn’t explosive. Benvenuto et al. [42] also found that COVID-2019 spread tends to reveal slightly decreasing spread. In addition, Heteroscedasticity (ARCH) were found to be insignificant in both models, indicating that the infection cases tend to show insignificants fluctuations. This is the fact that is not easily captured in the trends shown in Fig 12. Generally speaking, the diagnostic statistics indicate that the estimated models are acceptable since Q-statistics reveal that the residuals are not significantly correlated and the Jarque Berra statistic support the normality of residuals at conventional significance level. Also, ARCH effect was not significant, indicating a low volatility in the infection cases trend. In addition, all AR and MA roots were found to lie inside the unit circle, indicating that ARMA process is (covariance) stationary and invertible.

## Discussion

There is a great necessity for new robust scientific outcomes that could aid in containing and preventing the COVID-19 pandemic from spreading. The spatial mapping of COVID-19 outbreak risk can aid governments and policy-makers in implementing strict measures in certain regions of a city or a country where the risk of outbreak is very high. It is therefore crucial to identify the regions that would have high outbreak risk through predictive modelling with the help of machine learning algorithms (MLAs). In recent times, MLAs have demonstrated promising results in forecasting the epidemic outbreak risk [17]. In this research, the SVM model showing good forecast accuracy was used for mapping the outbreak risk of COVID-19. Similarly, Mohammadinia et al. [20] revealed that GWR and SVM had the highest precision in mapping the occurrence of leptospirosis. Ding et al. [47] employed three MLAs including SVM, RF and GBM for mapping the transmission risk assessment of mosquito-borne diseases and disclosed that all three MLAs acquired excellent validation outcome. Machado et al. [48] also applied RF, SVM and GBM in modelling the porcine epidemic diarrhoea virus and demonstrated 90% specificity values in case of SVM. Tien Bui et al. [17] stated that SVM achieved an AUC value of 0.968 in mapping the susceptibility to malaria. The ability to classify inseparable data classes is the greatest benefit of SVM model [49]. It is among the most precise and robust MLA [50]. SVM can be useful and has higher prediction accuracy when it comes to handling a small dataset. However, Huang and Zhao [51] demonstrated that SVM also yields excellent precision in predictive modelling when a large dataset is utilized. The algorithm have a very low probability to overfit and is not disproportionately impacted by noisy data [49]. Behzad et al. [52] revealed that SVM had huge capacity in simplification and had enduring forecast accuracy. It should be also noted that the predictive exactness of SVM model largely depends on the choice of kernel function [50]. Among the four kernel functions of SVM, RBF has been proved to generate high accuracy models [49]. SVM includes diverse kinds of categorization functions which are responsible for assessing overfitting and simplifying data that needs a minor tuning of model parameters [53]. The significance of each effective factor employed in this research is assessed using ridge regression. Since, there is no previous study in COVID-19 that outlines the proper effective factors. The outcome of this research can be very helpful for scientists in experimenting the same and additional effective factors for COVID-19 outbreak risk mapping. The proximity factors including distance from bus stations, distance from hospitals, distance from bakeries were most influential in forecasting the COVID-19 outbreak risk whereas other proximity factors such as distance from ATMs, distance from attraction sites, distance from fuel stations, distance from mosques and distance from road had the moderate influence which is followed by MTCM, density of cities and density of villages. It should be noted that climatic factors including MTWM, PWM and PDM had the least significance in mapping the outbreak risk. From this, it can be concluded that precipitation factors PWM and PDM are not associated with the transmission of COVID-19 in Fars Province whereas in case of temperature factors MTCM had moderate influence in mapping COVID-19 outbreak risk but MTWM exhibited a least significance. This outcome reveals that proximity factors had high influence in the transmission of SARS-CoV-2. In addition, the study conducted disclosed that increase in temperature will not decline the SARS-CoV-2 cases, although it has been also revealed that increase in temperature and absolute humidity could decrease the death of patients affected by COVID-19 [54]. A third-degree polynomial and ARMA models were applied to examine the behaviour of infection in Fars province and Iran. The general trend of infection in Iran and Fars province are similar while more explosive behavior for Iran’s cases is expected. The methodology and effective factors used in this research can be adapted in studies investigated in other parts of the world for preventing and controlling the outbreak risk of COVID-19.

## Conclusions

Mapping of SARS-CoV-2 outbreak risk can aid decision makers in drafting effective policies to minimize the spread of the disease. In this research, GIS based SVM was used for mapping the COVID-19 outbreak risk in Fars Province of Iran. Sixteen effective factors including MTCM, MTWM, PWM, PDM, distance from roads, distance from mosques, distance from hospitals, distance from fuel stations, human footprint, density of cities, distance from bus stations, distance from banks, distance from bakeries, distance from attraction sites, distance from automated teller machines (ATMs) and density of villages were selected along with the locations of active cases of SARS-CoV-2. The results of ridge regression revealed that distance from bus stations, distance from hospitals, and distance from bakeries had the highest significance and the outcome was utilized in mapping the outbreak risk of the pandemic with the help of SVM. The generated model had good predictive accuracy of 0.786 and 0.799 when verified with the locations of active cases during March 20 and March 29, 2020. The Iranian government should take restrict preventive measures for controlling the outbreak of SARS-CoV-2 in Shiraz as a tourism destination and the counties having high risk. Based on the results of polynomial and an ARMA model, the infection behavior is not expected to reveal an explosive process, however; the general trend of infection will last for several months especially in the Iran as a whole. A more slowly trend is expected in Fars Province, demonstrating extensive home quarantine and travel and movement restrictions were good strategies for disease control in Fars province. The main policy implication is that the infection cases, to some extent, may be controlled using more effective measures. Although, the estimated models may be used to predict the infection in following days, however; this contribution is less significant than the other implications derived from them. Generally speaking, it is expected to encounter a decreasing trend, however; this may be reversed if the ongoing attempts are slowed down, pointing out the need to keep the measures like quarantine or even to try more restricting attempts.

## Data Availability

All the data are available in the manuscript

## Availability of data and materials

All data and materials used in this work were publicly available.

## Ethics approval and consent to participate

The ethical approval or individual consent was not applicable

## Authors’ contributions

HRP, SP, BH, ZF, NS, MHT, BH, SB, and JPT contributed to study design, the literature search, data collection, data analysis, software working, and writing of this article. All authors read and approved the final draft of the manuscript.

## Consent for publication

Not applicable.

## References

1. Li, Q., Guan, X., Wu, P., Wang, X., Zhou, L., Tong, Y., Feng, Z. Early transmission dynamics in Wuhan, China, of Novel Coronavirus–Infected Pneumonia. New England Journal of Medicine. 2020. https://doi.org/10.1056/nejmoa2001316

2. Ma, Y., Zhao, Y., Liu, J., He, X., Wang, B., Fu, S., Luo, B. Effects of temperature variation and humidity on the death of COVID-19 in Wuhan, China. Science of the Total Environment. 2020; 138226. https://doi.org/10.1016/j.scitotenv.2020.138226

3. Huang, C., Wang, Y., Li, X., Ren, L., Zhao, J., Hu, Y., Cao, B. Clinical features of patients infected with 2019 novel coronavirus in Wuhan, China. The Lancet. 2020; 395(10223), 497–506. https://doi.org/10.1016/S0140-6736(20)30183-5

4. Wang, C., Horby, P. W., Hayden, F. G., Gao, G. F. A novel coronavirus outbreak of global health concern. The Lancet. 2020; 395, 470–473. https://doi.org/10.1016/S0140-6736(20)30185-9

5. Sohrabi, C., Alsafi, Z., O’Neill, N., Khan, M., Kerwan, A., Al-Jabir, A., Agha, R. World Health Organization declares global emergency: A review of the 2019 novel coronavirus (COVID-19). International Journal of Surgery. 2020; 76, 71–76. https://doi.org/10.1016/j.ijsu.2020.02.034

6. WHO, 2020a. WHO characterizes COVID-19 as a pandemic, 2020 (3).

7. WHO, 2020b. Coronavirus disease 2019 (COVID-19) Situation Report–70.

8. Remuzzi, A., Remuzzi, G. COVID-19 and Italy: what next? The Lancet. 2020; https://doi.org/10.1016/S0140-6736(20)30627-9

9. Arab-Mazar, Z., Sah, R., Rabaan, A. A., Dhama, K., Rodriguez-Morales, A. J. Mapping the incidence of the COVID-19 hotspot in Iran – Implications for Travellers. Travel Medicine and Infectious Disease. 2020; 101630. https://doi.org/10.1016/j.tmaid.2020.101630

10. Takian, A., Raoofi, A., Kazempour-Ardebili, S. COVID-19 battle during the toughest sanctions against Iran. Lancet (London, England). 2020; (20), 30668. https://doi.org/10.1016/S0140-6736(20)30668-1

11. Singh, A. K., Singh, A., Shaikh, A., Singh, R., Misra, A. Chloroquine and hydroxychloroquine in the treatment of COVID-19 with or without diabetes: A systematic search and a narrative review with a special reference to India and other developing countries. Diabetes & Metabolic Syndrome: Clinical Research & Reviews. 2020; https://doi.org/10.1016/j.dsx.2020.03.011

12. McCloskey, B., Zumla, A., Ippolito, G., Blumberg, L., Arbon, P., Cicero, A., Borodina, M. Mass gathering events and reducing further global spread of COVID-19: a political and public health dilemma. The Lancet. 2020; https://doi.org/10.1016/S0140-6736(20)30681-4

13. Zhou, C., Su, F., Pei, T., Zhang, A., Du, Y., Luo, B., Xiao, H. COVID-19: Challenges to GIS with Big Data. Geography and Sustainability. 2020; https://doi.org/10.1016/j.geosus.2020.03.005

14. Sánchez-Vizcaíno, F., Martínez-López, B., Sánchez-Vizcaíno, J. M. Identification of suitable areas for the occurrence of Rift Valley fever outbreaks in Spain using a multiple criteria decision framework. Veterinary Microbiology. 2013; 165(1–2), 71–78. https://doi.org/10.1016/j.vetmic.2013.03.016

15. Reeves, T., Samy, A. M., Peterson, A. T. MERS-CoV geography and ecology in the Middle East: Analyses of reported camel exposures and a preliminary risk map. BMC Research Notes. 2015; 8(1), 1–7. https://doi.org/10.1186/s13104-015-1789-1

16. Nyakarahuka, L., Ayebare, S., Mosomtai, G., Kankya, C., Lutwama, J., Mwiine, F. N., Skjerve, E. Ecological Niche Modeling for Filoviruses: A Risk Map for Ebola and Marburg Virus Disease Outbreaks in Uganda. PLoS Currents. 2017; 9. https://doi.org/10.1371/currents.outbreaks.07992a87522e1f229c7cb023270a2af1

17. Tien Bui, Q. T., Nguyen, Q. H., Pham, V. M., Pham, M. H., Tran, A. T. Understanding spatial variations of malaria in Vietnam using remotely sensed data integrated into GIS and machine learning classifiers. Geocarto International. 2019; 34(12), 1300–1314. https://doi.org/10.1080/10106049.2018.1478890

18. Jiang, D., Hao, M., Ding, F., Fu, J., & Li, M. Mapping the transmission risk of Zika virus using machine learning models. Acta Tropica. 2018; 185, 391–399. https://doi.org/10.1016/j.actatropica.2018.06.021

19. Carvajal, T. M., Viacrusis, K. M., Hernandez, L. F. T., Ho, H. T., Amalin, D. M., Watanabe, K. Machine learning methods reveal the temporal pattern of dengue incidence using meteorological factors in metropolitan Manila, Philippines. BMC Infectious Diseases. 2018; 18(1), 183. https://doi.org/10.1186/s12879-018-3066-0

20. Mohammadinia, A., Saeidian, B., Pradhan, B., Ghaemi, Z. Prediction mapping of human leptospirosis using ANN, GWR, SVM and GLM approaches. BMC Infectious Diseases. 2019; 19(1), 971. https://doi.org/10.1186/s12879-019-4580-4

21. Kamel Boulos, M. N., Geraghty, E. M. Geographical tracking and mapping of coronavirus disease COVID-19/severe acute respiratory syndrome coronavirus 2 (SARS- CoV-2) epidemic and associated events around the world: how 21st century GIS technologies are supporting the global fight against outbreaks and epidemics. International Journal of Health Geographics. 2020; 19, 8. https://doi.org/10.1186/s12942-020-00202-8

22. Chen, S., Yang, J., Yang, W., Wang, C., Bärnighausen, T. COVID-19 control in China during mass population movements at New Year. The Lancet. 2020; 395, 764–766. https://doi.org/10.1016/S0140-6736(20)30421-9

23. Wang, M., Jiang, A., Gong, L., Luo, L., Guo, W., Li, C., Li, H. Temperature significant change COVID-19 Transmission in 429 cities. MedRxiv. 2020; 20025791. https://doi.org/10.1101/2020.02.22.20025791

24. Tan, J., Mu, L., Huang, J., Yu, S., Chen, B., Yin, J. An initial investigation of the association between the SARS outbreak and weather: With the view of the environmental temperature and its variation. Journal of Epidemiology and Community Health. 2005; 59(3), 186–192. https://doi.org/10.1136/jech.2004.020180

25. Correa Ayram, C. A., Mendoza, M. E., Etter, A., Pérez Salicrup, D. R. Anthropogenic impact on habitat connectivity: A multidimensional human footprint index evaluated in a highly biodiverse landscape of Mexico. Ecological Indicators. 2017; 72, 895–909. https://doi.org/10.1016/j.ecolind.2016.09.007

26. Tarwater, P. M., Martin, C. F. Effects of population density on the spread of disease. Complexity. 2001; 6(6), 29–36. https://doi.org/10.1002/cplx.10003

27. Schmidt, W. P., Suzuki, M., Thiem, V., White, R. G., Tsuzuki, A., Yoshida, L. M., Ariyoshi, K. Population density, water supply, and the risk of dengue fever in Vietnam: Cohort study and spatial analysis. PLoS Medicine. 2011; 8(8). https://doi.org/10.1371/journal.pmed.1001082

28. Gilbert, M., Pullano, G., Pinotti, F., Valdano, E., Poletto, C., Boëlle, P. Y., Colizza, V. Preparedness and vulnerability of African countries against importations of COVID-19: a modelling study. The Lancet. 2020; 395(10227), 871–877. https://doi.org/10.1016/S0140-6736(20)30411-6

29. Pourghasemi, H. R., Kariminejad, N., Amiri, M., Edalat, M., Zarafshar, M., Blaschke, T., Cerda, A. Assessing and mapping multi-hazard risk susceptibility using a machine learning technique. Scientific Reports. 2020; 10(1), 3203. https://doi.org/10.1038/s41598-020-60191-3

30. Hoerl, A. E., Kennard, R. W. Ridge Regression: Biased Estimation for Nonorthogonal Problems. Technometrics. 1970; 12(1), 55–67. https://doi.org/10.1080/00401706.1970.10488634

31. Tikhonov, A. N., Goncharsky, A. V., Stepanov, V. V., Yagola, A. G., Tikhonov, A. N., Goncharsky, A. V., Yagola, A. G. Regularization methods. In Numerical Methods for the Solution of Ill-Posed Problems. 1995; 7–63. https://doi.org/10.1007/978-94-015-8480-7_2

32. Vapnik, V.N., An overview of statistical learning theory. IEEE Trans. Neural Network. 1999; 10 (5): 988–999. https://doi.org/10.1109/72.788640.

33. Gayen, A., Pourghasemi, H. R., Saha, S., Keesstra, S., Bai, S. Gully erosion susceptibility assessment and management of hazard-prone areas in India using different machine learning algorithms. Science of the Total Environment. 2019; 668, 124–138. https://doi.org/10.1016/j.scitotenv.2019.02.436

34. Yousefi, S., Sadhasivam, N., Pourghasemi, H. R., Ghaffari Nazarlou, H., Golkar, F., Tavangar, S., Santosh, M. Groundwater spring potential assessment using new ensemble data mining techniques. Measurement. 2020; 157, 107652. https://doi.org/10.1016/j.measurement.2020.107652

35. Pourghasemi, H. R., Sadhasivam, N., Kariminejad, N., Collins, A. Gully erosion spatial modelling: Role of machine learning algorithms in selection of the best controlling factors and modelling process. Geoscience Frontiers. 2020; https://doi.org/10.1016/j.gsf.2020.03.005

36. Garosi, Y., Sheklabadi, M., Conoscenti, C., Pourghasemi, H. R., Van Oost, K. Assessing the performance of GIS-based machine learning models with different accuracy measures for determining susceptibility to gully erosion. Science of the Total Environment. 2019; 664, 1117–1132. https://doi.org/10.1016/j.scitotenv.2019.02.093

37. Yao, X., Tham, L. G., Dai, F. C. Landslide susceptibility mapping based on Support Vector Machine: A case study on natural slopes of Hong Kong, China. Geomorphology. 2008; 101(4), 572–582. https://doi.org/10.1016/j.geomorph.2008.02.011

38. Chen, W., Pourghasemi, H. R., Naghibi, S. A. A comparative study of landslide susceptibility maps produced using support vector machine with different kernel functions and entropy data mining models in China. Bulletin of Engineering Geology and the Environment. 2018; 77(2), 647–664. https://doi.org/10.1007/s10064-017-1010-y

39. Dodangeh, E., Choubin, B., Eigdir, A. N., Nabipour, N., Panahi, M., Shamshirband, S., Mosavi, A. Integrated machine learning methods with resampling algorithms for flood susceptibility prediction. Science of the Total Environment. 2020; 705, 135983. https://doi.org/10.1016/j.scitotenv.2019.135983

40. Aik, J., Heywood, A. E., Newall, A. T., Ng, L.C., Kirk, M. D., Turner, R. Climate variability and salmonellosis in Singapore – A time series analysis. Science of the Total Environment. 2018; 639, 1261–1267.

41. Enders, W. Applied Econometric Times Series. John Wiley & Sons. 2004.

42. Benvenuto, D., Giovanetti, M., Vassallo, L., Angeletti, S., Ciccozzi, M. Application of the ARIMA model on the COVID-2019 epidemic dataset. Data in Brief. 2020; 29:105340.

43. Wong, G. Has SARS infected the property market? Evidence from Hong Kong. Journal of Urban Economics. 2008; 63, 74–95.

44. Alberts, C. J., Boyd, A., Bruisten, S. M., Heijman, T., Hogewoning, A., van Rooijen, M., Siedenburg, E., Sonder, G. J. B. Hepatitis A incidence, seroprevalence, and vaccination decision among MSM in Amsterdam, the Netherlands. Vaccine. 2019; 37, 2849–2856.

45. Leonenko, V. N., Ivanov, S. V., Novoselova, Y. K. A computational approach to investigate patterns of acute respiratory illness dynamics in the regions with distinct seasonal climate transitions. Procedia Computer Science. 2016; 80, 2402–2412.

47. Ding, F., Fu, J., Jiang, D., Hao, M., Lin, G. Mapping the spatial distribution of Aedes aegypti and Aedes albopictus. Acta Tropica. 2018; 178, 155–162. https://doi.org/10.1016/j.actatropica.2017.11.020

48. Machado, G., Vilalta, C., Recamonde-Mendoza, M., Corzo, C., Torremorell, M., Perez, A., VanderWaal, K. Identifying outbreaks of Porcine Epidemic Diarrhea virus through animal movements and spatial neighborhoods. Scientific Reports. 2019; 9(1), 1–12. https://doi.org/10.1038/s41598-018-36934-8

49. Gigovic, L., Pourghasemi, H. R., Drobnjak, S., Bai, S. Testing a New Ensemble Model Based on SVM and Random Forest in Forest Fire Susceptibility Assessment and Its Mapping in Serbia’s Tara National Park. Forests. 2019; 10(5), 408. https://doi.org/10.3390/f10050408

50. Abdollahi, S., Pourghasemi, H. R., Ghanbarian, G. A., Safaeian, R. Prioritization of effective factors in the occurrence of land subsidence and its susceptibility mapping using an SVM model and their different kernel functions. Bulletin of Engineering Geology and the Environment. 2019; 78(6), 4017–4034. https://doi.org/10.1007/s10064-018-1403-6

51. Huang, Y., Zhao, L. Review on landslide susceptibility mapping using support vector machines. Catena. 2018; 165: 520–529. https://doi.org/10.1016/j.catena.2018.03.003

52. Behzad, M., Asghari, K., Coppola, E. A. Comparative Study of SVMs and ANNs in Aquifer Water Level Prediction. Journal of Computing in Civil Engineering. 2010; 24(5): 408–413. https://doi.org/10.1061/(ASCE)CP.1943-5487.0000043

53. Joachims, T. Text categorization with support vector machines: Learning with many Relevant Features. 1998. https://doi.org/10.1007/bfb0026683

54. Zhu, Y., Xie, J. Association between ambient temperature and COVID-19 infection in 122 cities from China. Science of the Total Environment, 2020; 138201. https://doi.org/10.1016/j.scitotenv.2020.138201

